# Neonatal critical illness is associated with pancytopenia development in childhood

**DOI:** 10.64898/2026.07.21.26358620

**Authors:** Brian M. Dulmovits, Nicolas P. Goldstein Novick, Matthew Devine, Christopher S. Thom

## Abstract

**Objectives:** Perinatal illness and preterm birth carry lifelong multiorgan complications and are associated with hematologic derangements during neonatal intensive care unit (NICU) admission. Despite this, long term hematologic morbidities following neonatal critical illness remain undefined. Our objective was to identify associations between prematurity, perinatal critical illness, and later hematologic dysfunction.

**Study design:** Single neonatal care network retrospective cohort study with cohorts divided by gestational age and the presence of critical illness markers. The association between hematologic dysfunction, critical illness, and prematurity was investigated using multivariate logistic regression.

**Results:** Among 13073 infants, critical illness or prematurity was found to increase the odds of developing pancytopenia post-NICU discharge. Subsequent analyses stratified on prematurity demonstrate that a diagnosis of shock or sepsis was associated with pancytopenia.

**Conclusions:** Our findings suggest that perinatal insults are associated with hematopoietic system dysfunction and long term morbidity. Importantly, critical illness, not prematurity itself, may drive this association in preterm infants.

## Introduction

Premature birth and perinatal illness can be acutely life-threatening and cause long term morbidities. For example, preterm birth increases risks of cardiometabolic syndromes, impaired lung function, neurodevelopment impairment, and prolonged growth failure.^1–6^ Neonatal sepsis impacts long-term neurodevelopment in term and preterm infants.^7^ Critical illness requiring extracorporeal membrane oxygenation (ECMO) adversely affects neurodevelopment, renal and pulmonary function.^8^ With increased survivorship among extremely preterm and critically ill neonates, defining long term complications of perinatal illness have taken on increased importance.

Few studies have examined long term hematologic complications associated with perinatal illness or prematurity. Many infants hospitalized in the neonatal intensive care unit (NICU) experience quantitative blood count abnormalities, including elevated (thrombocytosis, polycythemia, leukocytosis) or low blood counts (anemia, thrombocytopenia, leukopenia).^9–12^ Cytopenias typically occur secondary to another inciting condition, such as sepsis, thrombosis, or iatrogenic blood loss from phlebotomy. Clinical responses therefore focus on treatment of the underlying pathology, as well as blood product transfusions to acutely replenish deficient blood cells.^13^ However, recurrent cytopenias and quantitative blood count variation could also result from immaturity or injury to the bone marrow as the primary hematopoietic organ. For example, reversible pancytopenia can be observed among toddlers and adolescents hospitalized in the pediatric intensive care unit (PICU) with critical illness. While case reports exist, there has not been a formal analysis of perinatal events that might underlie a predisposition to develop short-term pancytopenia during stress in later childhood.

Bone marrow insult can compromise hematopoietic output, particularly during times of stress (*e.g*., infection and inflammation), resulting in pancytopenia. This is sometimes referred to clinically as having ‘low bone marrow reserve’ of hematopoietic stem and progenitor cells (HSPC), a process akin to acute bone marrow failure. Inflammation and bone marrow stress can also promote clonal hematopoiesis and leukemic transformation.^14,15^ Links between preterm birth, low birth weight, mechanical ventilation, and later development of leukemia and clonal hematopoiesis suggest that perinatal insults may produce enduring injury to bone marrow hematopoiesis.^16–18^ In extremely preterm infants, critical illness may affect bone marrow as it undergoes HSPC colonization.

Few studies have investigated nonmalignant hematopoietic morbidities following preterm birth and/or neonatal illness. Here, we sought to identify associations between perinatal insult among term and preterm infants, and the development of later childhood hematologic sequelae, in a large pediatric cohort.

## Methods

### Study design and population

We performed a single care network retrospective cohort study using electronic health record (EHR) data derived from participants cared for at the Children’s Hospital of Philadelphia (CHOP). We included any infant admitted to the level IV CHOP neonatal/infant intensive care unit (N/IICU) and associated level III NICU, as well as infants with outpatient encounters at CHOP associated Neonatal Follow-up Programs, from 2008-2025. This study was approved by the Children’s Hospital of Philadelphia Institutional Review Board (IRB 26-024728).

### Primary and secondary outcomes

Our primary outcome was a composite measure of hematologic dysfunction, defined as any diagnosis of pancytopenia, bone marrow failure, myelodysplasia, or leukemia. Secondary outcomes included each individual hematologic diagnosis. We identified hematologic dysfunction using International Classification of Diseases (ICD) diagnostic codes, which relied on participants returning for follow up care within the CHOP Care Network. If pancytopenia and another hematologic disorder were diagnosed within one month of each other, the other hematologic disorder was utilized as the primary diagnosis. If multiple diagnoses other than pancytopenia were diagnosed in the same child, we utilized the earlier date to define the date of hematologic dysfunction. We excluded from analysis participants with missing data. We also excluded participants with diagnoses of congenital leukemia or Trisomy 21 to avoid confounding from blood cancer predisposition syndromes.

### Primary exposure

We defined our primary exposure as the combination of prematurity and neonatal critical illness. We first categorized infants as term (gestational age [GA] ≥37 weeks) or preterm (GA <37 weeks). We then further classified term infants according to predefined indicators of critical illness (**Fig 1**; **Table 1**), generating non-critically ill term (control), critically ill term, and preterm groups. We obtained medication exposures, respiratory support, and laboratory and clinical diagnoses (e.g., positive blood culture) from the EHR. We defined vasoactive medication exposure as dopamine, epinephrine, norepinephrine, phenylephrine, vasopressin, or hydrocortisone; systemic steroid exposure as dexamethasone, methylprednisolone, prednisone, or prednisolone; and non-invasive ventilation as continuous positive airway pressure, bilevel positive airway pressure, high flow nasal cannula, or > 2L nasal cannula. We defined the control group as term infants admitted with cleft lip/palate, myelomeningocele, ventriculomegaly, and/or hyperbilirubinemia who had none of the predefined indicators of critical illness. We allowed infants with prematurity or critical illness indicators to also have one or more of these diagnoses.

**Figure 1.**
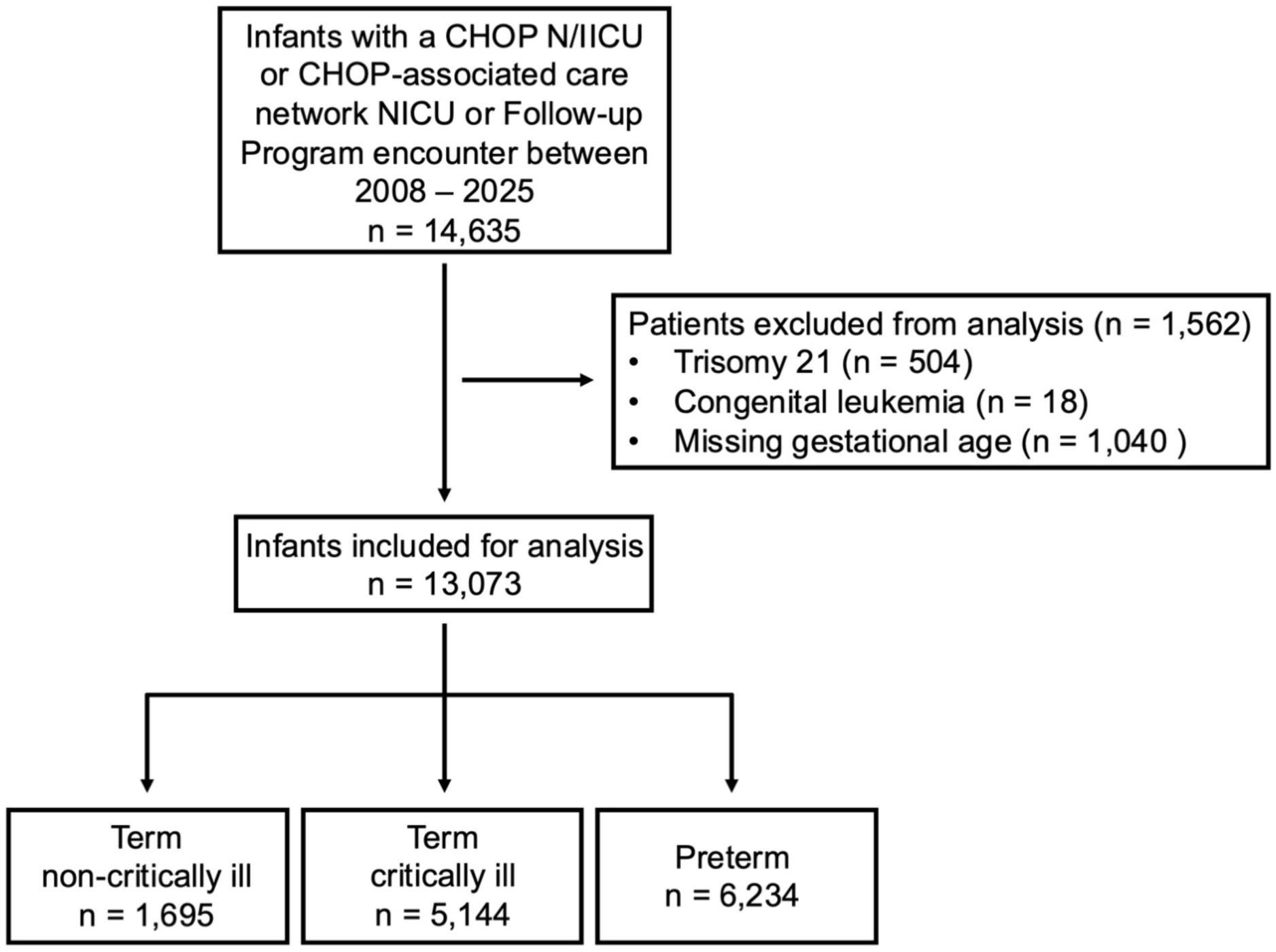
Flow diagram of study population.

**Table 1.** Demographics, length of stay, and indicators of critical illness and non-critical illness.

| N = 13,073 | Non-critically ill term<br>n = 1,695<br>mean (SD) or n (%) | Critically ill term<br>n = 5,144<br>mean (SD) or n (%) | Preterm<br>n = 6,234<br>mean (SD) or n (%) | P-value |
| --- | --- | --- | --- | --- |
| Year of Birth |  |  |  |  |
| 2008-2013 | 522 (30.8%) | 1,331 (25.9%) | 1,892 (30.4%) | <0.001<sup>A</sup> |
| 2014-2019 | 596 (35.2%) | 1,906 (37.1%) | 2,281 (36.6%) |  |
| 2020-2025 | 577 (34.0%) | 1,907 (37.1%) | 2,061 (33.1%) |  |
| Gestational Age (weeks)<br>Median (IQR) | 38.7 (37.6-39.3) | 39.0 (37.9-38.6) | 32.1 (27.1-35.0) | <0.001<sup>B</sup> |
| Birthweight (kg)<br>Median (IQR) | 3.2 (2.9-3.5) | 3.2 (2.9-3.6) | 1.6 (0.9-2.4) | <0.001<sup>B</sup> |
| Female Sex | 753 (44.4%) | 2,203 (42.8%) | 2,719 (43.6%) | 0.460<sup>A</sup> |
| Race |  |  |  |  |
| Asian | 107 (6.3%) | 227 (4.4%) | 201 (3.2%) | <0.001<sup>A</sup> |
| Black | 347 (20.5%) | 923 (18.0%) | 1,723 (27.7%) |  |
| Multi-Racial | 100 (5.9%) | 314 (6.1%) | 339 (5.5%) |  |
| Other | 246 (14.5%) | 915 (17.8%) | 1,071 (17.2%) |  |
| White | 878 (51.8%) | 2,653 (51.7%) | 2,772 (44.6%) |  |
| Unknown | 16 (0.9%) | 100 (2.0%) | 107 (1.7%) |  |
| Ethnicity |  |  |  |  |
| Hispanic | 191 (11.6%) | 714 (14.2%) | 837 (13.8%) | 0.023<sup>A</sup> |
| Non-Hispanic | 1,408 (85.2%) | 4,171 (82.8%) | 5,065 (83.7%) |  |
| Unknown | 53 (3.2%) | 153 (3.0%) | 151 (2.5%) |  |
| Length of Stay (days)<br>Median (IQR) | 4 (2-8) | 16 (7-35) | 34 (12-95) | <0.001<sup>B</sup> |
| Indicators of critical illness |  |  |  |  |
| Any vasoactives | 0 (0.0%) | 2,284 (44.4%) | 3,308 (53.1%) | <0.001<sup>C</sup> |
| Any invasive ventilation | 0 (0.0%) | 1,880 (40.7%) | 2,662 (49.1%) | <0.001<sup>C</sup> |
| Any non-invasive ventilation | 0 (0.0%) | 3,113 (67.4%) | 3,326 (61.4%) | <0.001<sup>C</sup> |
| Any systemic steroids | 0 (0.0%) | 1,040 (20.2%) | 1,494 (24.0%) | <0.001<sup>C</sup> |
| Any positive blood culture | 0 (0.0%) | 635 (12.3%) | 1,349 (21.6%) | <0.001<sup>C</sup> |
| Any sepsis | 0 (0.0%) | 1,329 (25.8%) | 1,736 (27.9%) | <0.001<sup>C</sup> |
| Any shock | 0 (0.0%) | 329 (6.4%) | 357 (5.7%) | <0.001<sup>C</sup> |
| ECMO | 0 (0.0%) | 383 (7.5%) | 132 (2.1%) | <0.001<sup>C</sup> |
| Tracheostomy | 0 (0.0%) | 117 (2.3%) | 360 (5.8%) | <0.001<sup>C</sup> |
| NEC | 0 (0.0%) | 97 (1.9%) | 639 (10.3%) | <0.001<sup>C</sup> |
| BPD | 0 (0.0%) | 0 (0.0%) | 1,387 (22.3%) | <0.001<sup>C</sup> |
| Grade III/IV IVH | 0 (0.0%) | 73 (1.4%) | 665 (10.7%) | <0.001<sup>C</sup> |
| PPHN/MAS | 0 (0.0%) | 1,123 (21.8%) | 536 (8.6%) | <0.001<sup>C</sup> |
| HIE | 0 (0.0%) | 1,115 (21.7%) | 355 (5.7%) | <0.001<sup>C</sup> |
| Indicators of non-critical illness |  |  |  |  |
| Cleft lip/palate | 92 (5.4%) | 310 (6.0%) | 193 (3.1%) | <0.001<sup>C</sup> |
| MMC | 211 (12.5%) | 298 (5.8%) | 385 (6.2%) | <0.001<sup>C</sup> |
| Ventriculomegaly | 436 (25.7%) | 1,535 (29.8%) | 2,660 (42.7%) | <0.001<sup>C</sup> |
| Hyperbilirubinemia | 1,035 (61.1%) | 1,346 (26.2%) | 2,563 (41.1%) | <0.001<sup>C</sup> |

Missingness: birthweight (799), female sex (3), race (34), ethnicity (330), length of stay (136), any invasive ventilation (1,501), any non-invasive ventilation (1,501)
<sup>A</sup>Chi-square test, <sup>B</sup>Kruskal-Wallis Test, <sup>C</sup>Fisher's exact test

### Descriptive variables

For each participant, as available, we collected the year of birth, gestational age (GA), birthweight, sex, race, ethnicity, and length of stay.

### Statistical analyses

We first compared year of birth, sociodemographic and medical characteristics, length of stay, indicators of critical illness, hematologic indices (white blood cell count, hemoglobin, and platelet count), conjugated and unconjugated bilirubin concentrations, and the frequency of hematologic dysfunction and its component diagnoses across non-critically ill term infants, critically ill term infants, and preterm infants using chi-square, Kruskal-Wallis, or Fisher’s exact tests, as appropriate. We then used logistic regression models with birth-year fixed effects to estimate associations of the prematurity-critical illness groups with hematologic dysfunction and its component diagnoses. For hematologic dysfunction and component diagnoses with significant overall associations, we performed exploratory analyses separately examining diagnoses during the birth hospitalization and after NICU discharge to evaluate the timing of associations. Finally, we used logistic regression models with birth-year fixed-effects, stratified by prematurity status, to evaluate associations between individual markers of critical illness and hematologic dysfunction or component diagnoses with significant overall associations among term and preterm infants separately, using unadjusted and multivariable models that included all critical illness markers.

## Results

### Identification of critically ill and premature infant cohorts

We identified 13,073 participants who met inclusion criteria, 48% of whom were born preterm (mean GA 32.1 weeks, **Table 1**). Of preterm infants, 28% were born 22-28 weeks’ gestation. Full term infants with at least one marker of critical illness represented 39% of the cohort (**Table 1**). Non-invasive ventilation (67% of critically ill term and 61% preterm infants), vasoactive requirement (44% and 53%), and invasive ventilation (41% and 49%) were the most frequently observed indicators of critical illness (**Table 1**). Sepsis was diagnosed in 26% and 28% of critically ill term and preterm infants, respectively. The development of critical illness markers was variable in preterm infants occurring in 2 – 61% of this cohort and included NEC, BPD, vasoactive medications, and invasive ventilation. The elevated proportion of preterm infants with critical illness markers reflects the elevated severity of illness within our preterm cohort requiring level IV NICU admission.

Compared to non-critically ill term infants, critically ill term and preterm infants showed differences in hematologic indices and bilirubin concentrations (**Fig. 2A; Supplemental Table 1**) and exhibited higher incidences of composite hematologic dysfunction (**Fig. 2B; Supplemental Table 2**). Pancytopenia was the most frequently observed hematologic diagnosis among those collected, and was significantly increased among preterm and critically ill cohorts compared to controls (**Fig. 2B; Supplemental Table 2**). Unconjugated bilirubin levels were significantly increased among full term control participants, consistent with the intuition that many of those individuals were admitted for management of hyperbilirubinemia (**Table 1** and **Fig. 2A; Supplemental Table 1**).

**Figure 2.**
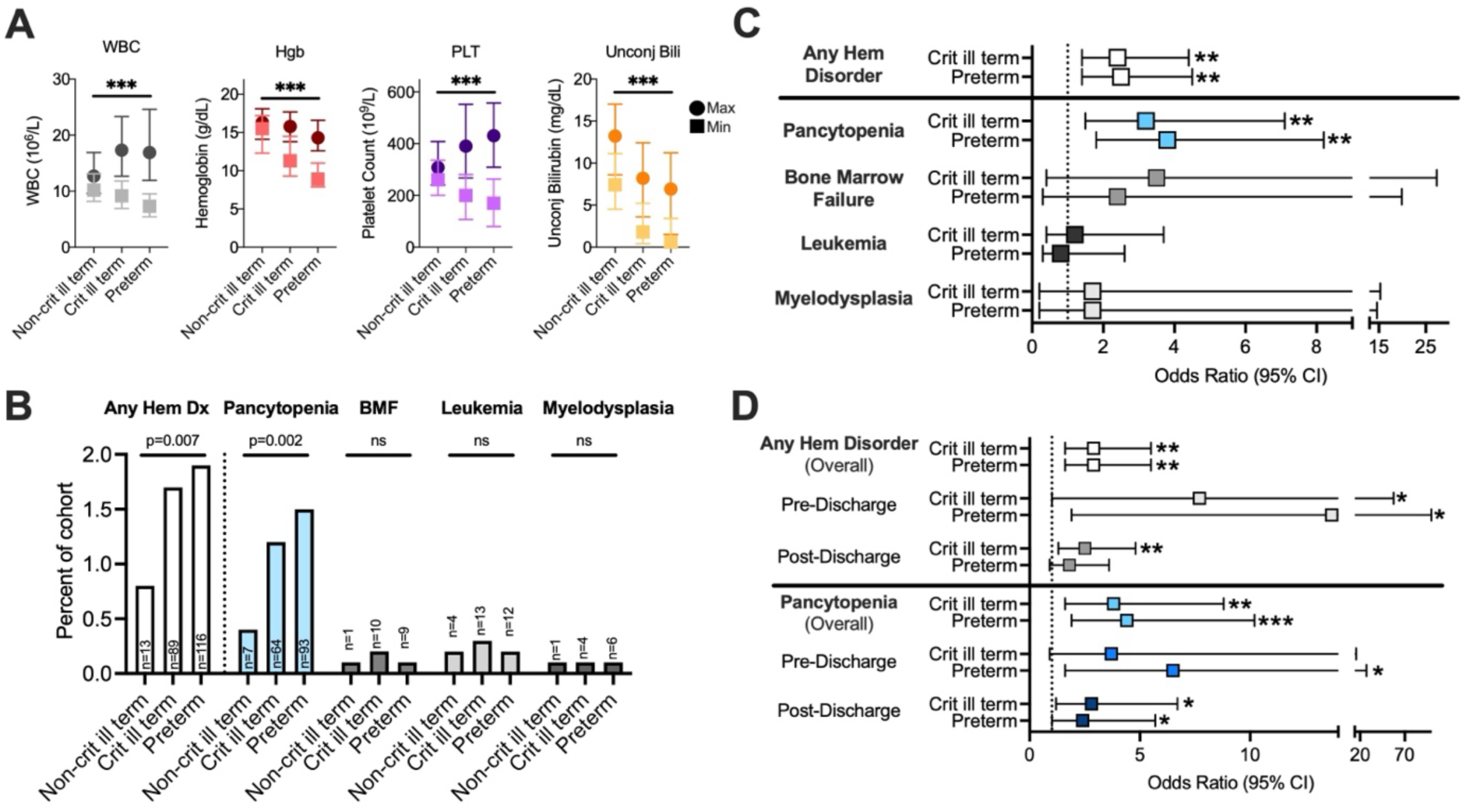
Hematologic disorders are linked to perinatal illness and/or premature birth. A. Complete blood count and bilirubin laboratory value variation among cohorts. Statistical significance assessed using Kruskal-Wallis test. B. Hematologic outcomes for each cohort. Statistical significance assessed using Chi-square test. C. Hematologic outcome prediction per cohort. Model reflects logistic regression with year fixed effects. Odds ratios with 95% confidence intervals. D. Hematologic disorders or pancytopenia diagnoses per cohort based on overall diagnosis, diagnosis pre-discharge, or post-discharge from the NICU. Model reflects logistic regression with year fixed effects. Odds ratios with 95% confidence intervals. *** p < 0.001, ** p < 0.01, * p < 0.05

### Premature birth and critical perinatal illness increase hematologic dysfunction risks

Logistic regression revealed increased odds of any diagnosis of hematologic dysfunction among term critical illness and preterm cohorts compared to controls (**Fig. 2C; Supplemental Table 3**). This association was driven by pancytopenia diagnosis. Odds ratios and incidence of bone marrow failure, leukemia, and myelodysplasia were increased among critically ill and/or preterm infants, but did not reach statistical significance (**Fig. 2B-C; Supplemental Table 3**).

To determine if pancytopenia or other hematologic derangements occurred during perinatal NICU admission and/or later in childhood, we stratified analyses to pre- and post-NICU discharge. Critically ill term and preterm infants both showed increased odds of pancytopenia post-discharge compared to controls (**Fig. 2D; Supplemental Table 4**). Preterm infants had greater odds of developing pancytopenia during NICU admission compared to controls, but critically ill term infants had no statistically significant difference. Of note, the preterm cohort demonstrated a trend towards earlier date of diagnosis (**Fig. 3**). This suggests that hematologic dysfunction associated with premature birth and/or perinatal illness may persist into childhood.

**Figure 3.**
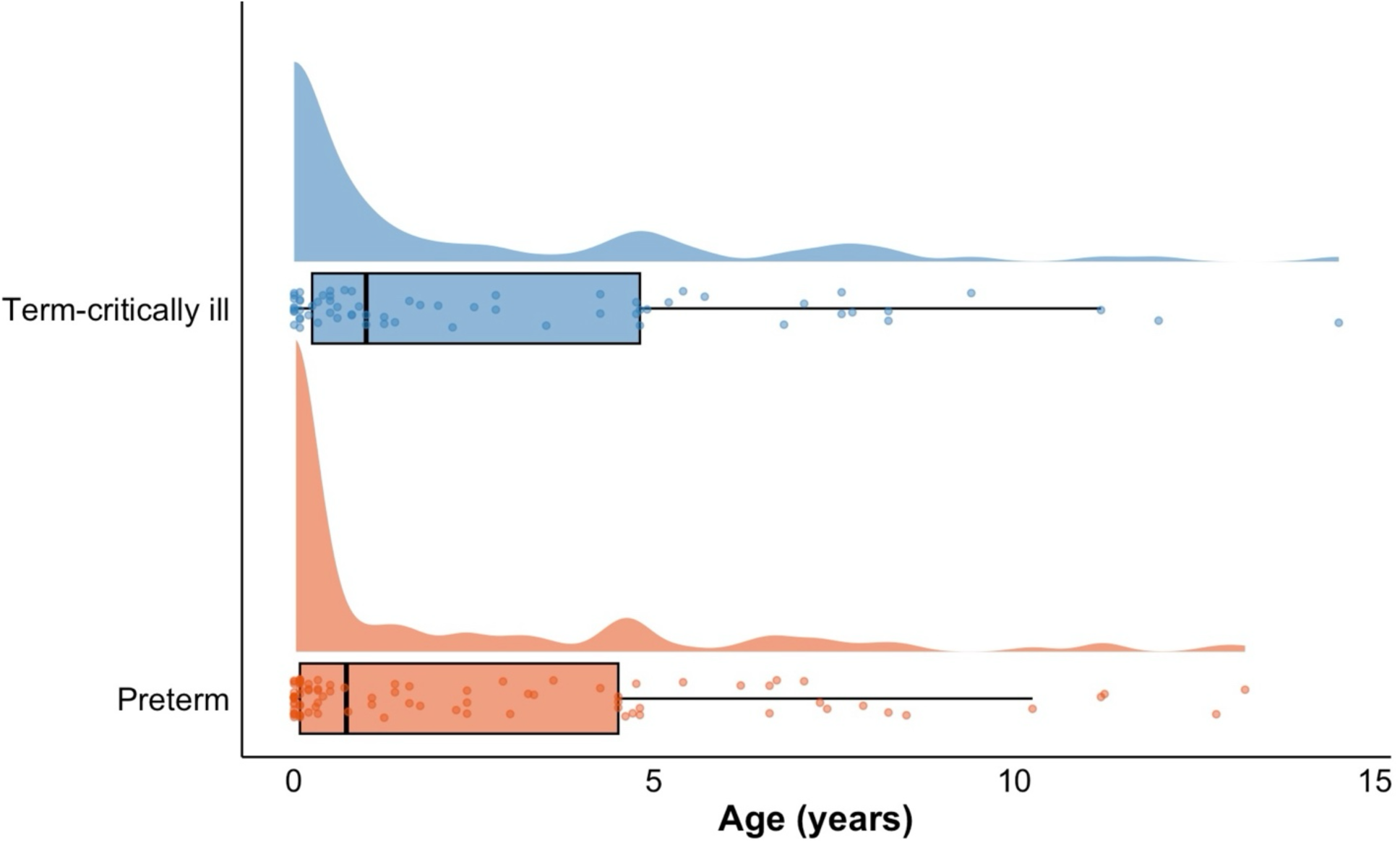
Distribution of pancytopenia cases by age of diagnosis. Combined density plots and boxplots of pancytopenia cases displayed by age of diagnosis in term-critically ill and preterm cohorts. Boxplots span the interquartile range (IQR) with the vertical black line indicating the median age of diagnosis and whiskers representing ± 1.5 x IQR.

### Prematurity and septic shock are associated with later hematologic dysfunction

We next wanted to identify the specific markers of perinatal illness that are associated with the development of pancytopenia or other hematologic disorders. We used unadjusted logistic regression with birth-year fixed effects stratified on prematurity to identify associations of individual critical illness markers with overall and post-discharge hematologic disorders or pancytopenia. Several markers of critical illness were associated with increased odds of hematologic disorders or a specific pancytopenia diagnosis in term critically ill and preterm infants (e.g., vasoactive medication use, invasive and non-invasive ventilation requirement, systemic steroid exposure, positive blood culture, sepsis, shock and NEC; **Fig. 4A-C; Supplemental Tables 5-7**). This was consistent with the hypothesis that critical illness in early life may alter bone marrow construction and/or function long term.

**Figure 4.**
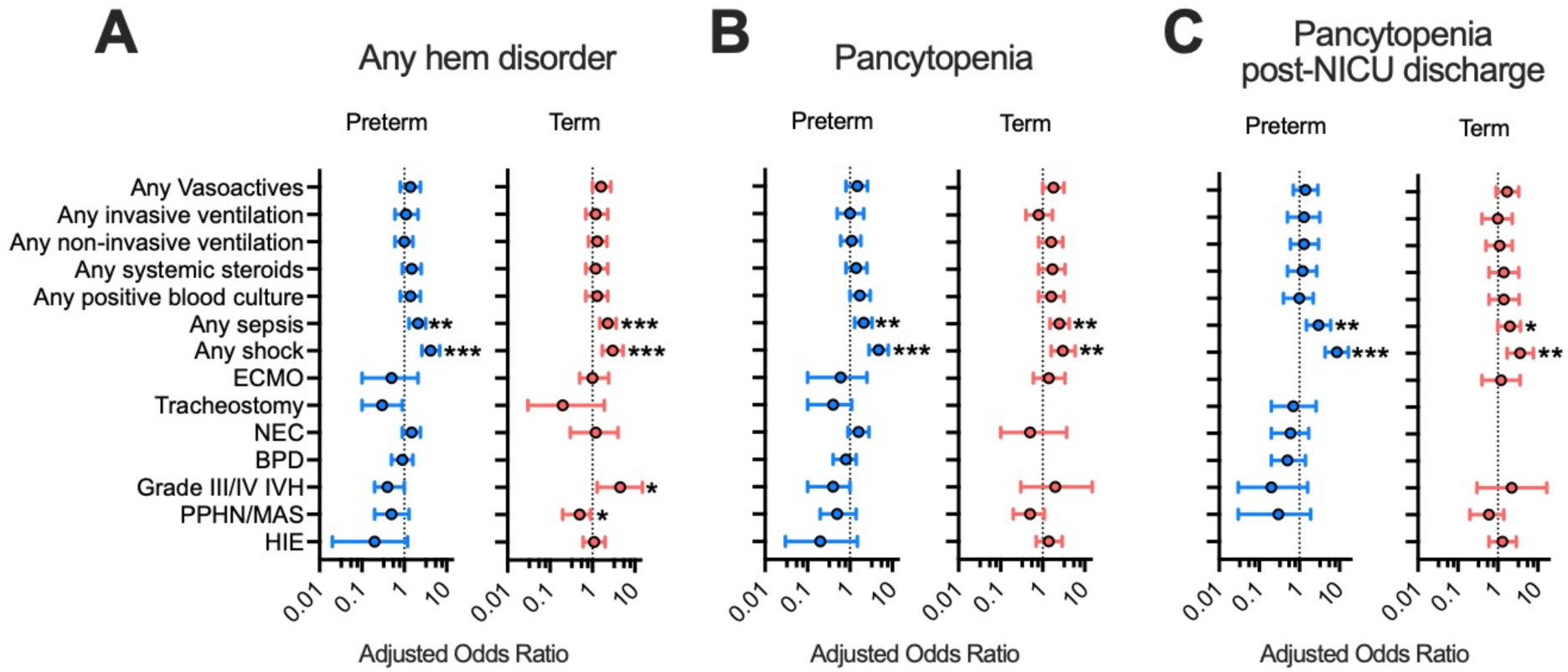
Key components of critical illness, including sepsis and shock, predict development of hematologic disorders. A. Multivariate adjusted odds ratios for individual components of critical illness in predicting any hematologic disorder, stratified on prematurity. B. Multivariate adjusted odds ratios for individual components of critical illness in predicting pancytopenia diagnosis, stratified on prematurity. C. Multivariate adjusted odds ratios for individual components of critical illness in predicting a diagnosis of pancytopenia post-discharge, stratified on prematurity. Models were built using logistic regression with year fixed effects, adjusted for displayed factors and additionally any non-invasive ventilation, extracorporeal membrane oxygenation, bronchopulmonary dysplasia, and hypoxic ischemic encephalopathy. *** p < 0.001, ** p < 0.01, * p < 0.05

We then wanted to untangle the associations controlling for all measured indicators of critical illness. Upon multivariable analysis stratified on prematurity, the effects of sepsis and shock were still robustly associated with hematologic disorder and pancytopenia overall and post-discharge (**Fig. 4A-C; Supplemental Tables 5-7**). This suggests that pathophysiology related to sepsis and shock may be the most important in driving these associations.

## Discussion

An association between neonatal critical illness and hematologic dysfunction in later childhood, specifically recurrent pancytopenia, suggests bone marrow dysfunction may be a consequence of perinatal insult. Premature birth and low birth weight have been linked to later development of leukemia and clonal hematopoiesis, which can also be a manifestation of bone marrow stress.^16–18^ Our findings extend this inferred bone marrow dysfunction to also be associated with critical illness in full term babies. Specifically, we found an association of pancytopenia after NICU discharge with neonatal sepsis and shock. We hypothesize that related mechanisms may trace to the initial construction of the bone marrow niche, which occurs in late gestation and supports hematopoiesis throughout life.

Early life infection alters the developmental trajectory of multiple organ compartments, including neurologic, ophthalmologic, pulmonary, cardiometabolic, and immune systems.^7,19–22^ Our study is, to our knowledge, the first to suggest novel long-term hematopoietic associations with neonatal infection in a clinical cohort. Mouse and human sepsis challenge models provide mechanistic explanation for these findings. Epigenetic changes in hematopoietic stem and progenitor cells perturb hematopoiesis following infection.^23–25^ Inflammatory conditions can also lead to the emergence of a long-lasting and distinct transcriptional state in human HSPCs.^26^ Polyinosinic:polycytidylic acid (PI:PC) treatment also induces changes to the bone marrow microenvironment in adult mice.^27^ Neonatal infection may also disrupt the initial construction of the hematopoietic niche, specifically stromal and mesenchymal cell populations that emerge in perinatal life.^28,29^

More broadly, our findings indicate that shock physiology is associated with later pancytopenia diagnosis and presumed hematopoietic dysfunction. We identified shock and vasoactive exposure as markers of critical illness associated with overall and post-discharge pancytopenia diagnoses. This would explain the discordance between the number of sepsis cases vs positive blood cultures in our cohort. Severe inflammatory responses, including “culture-negative sepsis” that mimics a sepsis phenotype, appears to impact hematopoietic system development and/or later function. Circulatory failure may lead to prolonged oxygen and nutrient deprivation in the bone marrow, beyond its normal state of relative hypoxia. Moreover, activation of adrenergic and renin-angiotensin-aldosterone systems through endogenous and exogenous (*e.g.* vasopressor infusions) during shock may alter hematopoiesis, as previously shown.^30–33^

The incidence of hematologic dysfunction was low in our study, including 218 cases over 17 years. Pancytopenia diagnoses were also rare, with an incidence of 1% (n = 64) and 2% (n = 93) in term critically ill and preterm groups, respectively. These numbers are significantly lower than isolated cytopenias frequently encountered during NICU admission, but higher than the incidence of other pediatric hematologic and oncologic conditions with high morbidity (e.g., immune-mediated aplastic anemia and leukemia).^34,35^ We envision that cases of long-term hematologic dysfunction associated with perinatal illness may rise with increased survivors of extreme prematurity and perinatal critical illness. Understanding which individuals may be at heightened risk for future hematologic pathology will inform longitudinal surveillance programs.

Our retrospective cohort was based on longitudinal EHR data from a single large neonatal care network. Patient-level data allowed greater resolution of perinatal insults compared to prior registries or meta-analysis studies, and facilitated associational analyses linking individual markers of critical illness with hematologic morbidities. However, using data from a single neonatal care network has inherent limitations. Most of the preterm cohort was derived from level IV NICU encounters, meaning that the population of premature infants was critically ill requiring transfer to our quaternary referral center. This likely affects the generalizability of our study for preterm infants who remain in birth hospitals. Identification of post-discharge hematologic dysfunction also relied on individuals receiving follow-up subspecialty care within our pediatric care network. This likely precluded detection of all cases of hematologic dysfunction, so we may have underestimated the incidence of some disorders. Given our study design, we are only able to characterize associations, which may be explained by unmeasured confounders.

Together, our study suggests that neonatal critical illness is associated with long-term hematopoietic dysfunction. This is a novel association that adds to the growing evidence that neonatal insults, including prematurity, may have with lifelong impacts. Our findings suggest that perinatal critical illness may need to be considered a risk factor when examining the hematologic sequelae of preterm birth. These findings warrant larger, multi-center studies with longer follow-up time and increased power to better detect rare events such as leukemia, myelodysplasia, and clonal hematopoiesis which may impact clinical care for neonatal, pediatric, and adult survivors of prematurity and perinatal illness.

## Acknowledgments

This study was supported by the National Institutes of Health (T32HL007150-47-49 to BMD, R00HL177827 to CST) and the Children’s Hospital of Philadelphia Division of Neonatology.

## Data Availability

The datasets generated and analyzed for this study are available upon request from the corresponding authors following completion of a data use agreement with the Children’s Hospital of Philadelphia.

## Supplemental Tables

**Supplemental Table 1.**
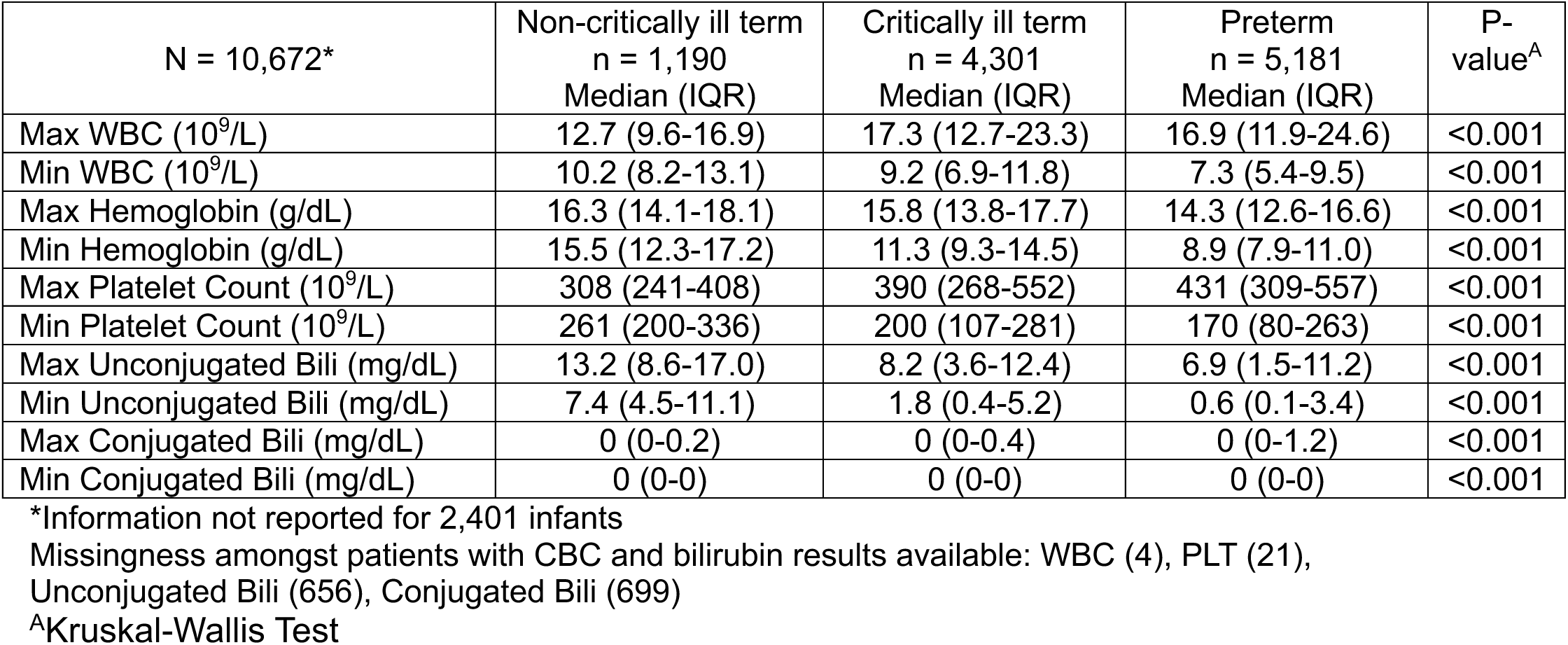
Select hematologic indices and bilirubin concentrations.

**Supplemental Table 2.**
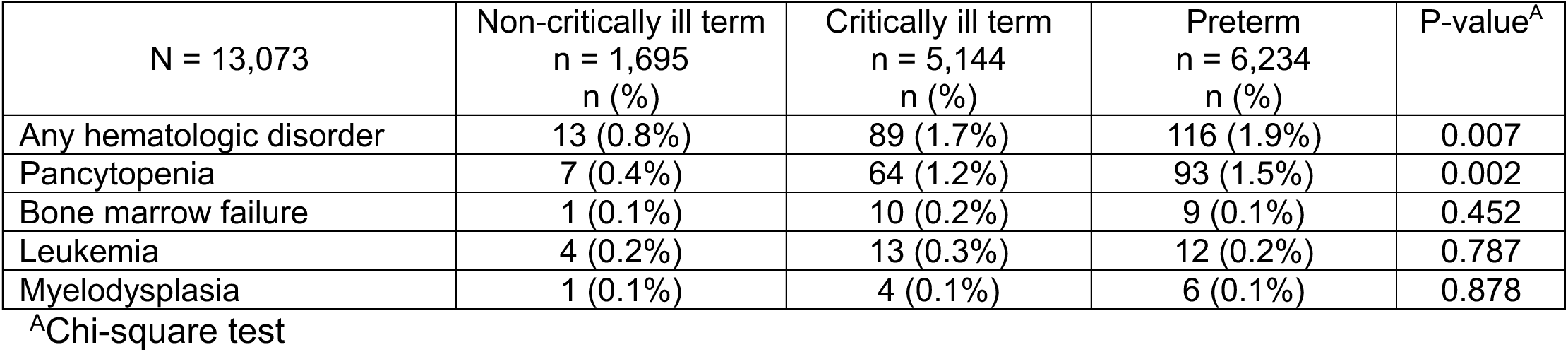
Counts of hematologic outcomes.

| N = 13,073 | Non-critically ill term<br>n = 1,695<br>n (%) | Critically ill term<br>n = 5,144<br>n (%) | Preterm<br>n = 6,234<br>n (%) | P-value<sup>A</sup> |
| --- | --- | --- | --- | --- |
| Any hematologic disorder | 13 (0.8%) | 89 (1.7%) | 116 (1.9%) | 0.007 |
| Pancytopenia | 7 (0.4%) | 64 (1.2%) | 93 (1.5%) | 0.002 |
| Bone marrow failure | 1 (0.1%) | 10 (0.2%) | 9 (0.1%) | 0.452 |
| Leukemia | 4 (0.2%) | 13 (0.3%) | 12 (0.2%) | 0.787 |
| Myelodysplasia | 1 (0.1%) | 4 (0.1%) | 6 (0.1%) | 0.878 |

<sup>A</sup>Chi-square test

**Supplemental Table 3.** Logistic regression predicting hematologic outcomes.

| N = 13,073 | Non-critically ill term<br>n = 1,696<br>OR (95% CI) | Critically ill term<br>n = 5,152<br>OR (95% CI) | Preterm<br>n = 6,235<br>OR (95% CI) |
| --- | --- | --- | --- |
| Any hematologic disorder | Reference | 2.4 (1.4-4.4)** | 2.5 (1.4-4.5)** |
| Pancytopenia | Reference | 3.2 (1.5-7.1)** | 3.8 (1.8-8.2)** |
| Bone marrow failure | Reference | 3.5 (0.4-27.3) | 2.4 (0.3-18.9) |
| Leukemia | Reference | 1.2 (0.4-3.7) | 0.8 (0.3-2.6) |
| Myelodysplasia | Reference | 1.7 (0.2-15.3) | 1.7 (0.2-14.6) |

\*\*\* <0.001, \*\* <0.01, \* <0.05; Model = logistic regression with year fixed effects

**Supplemental Table 4.**
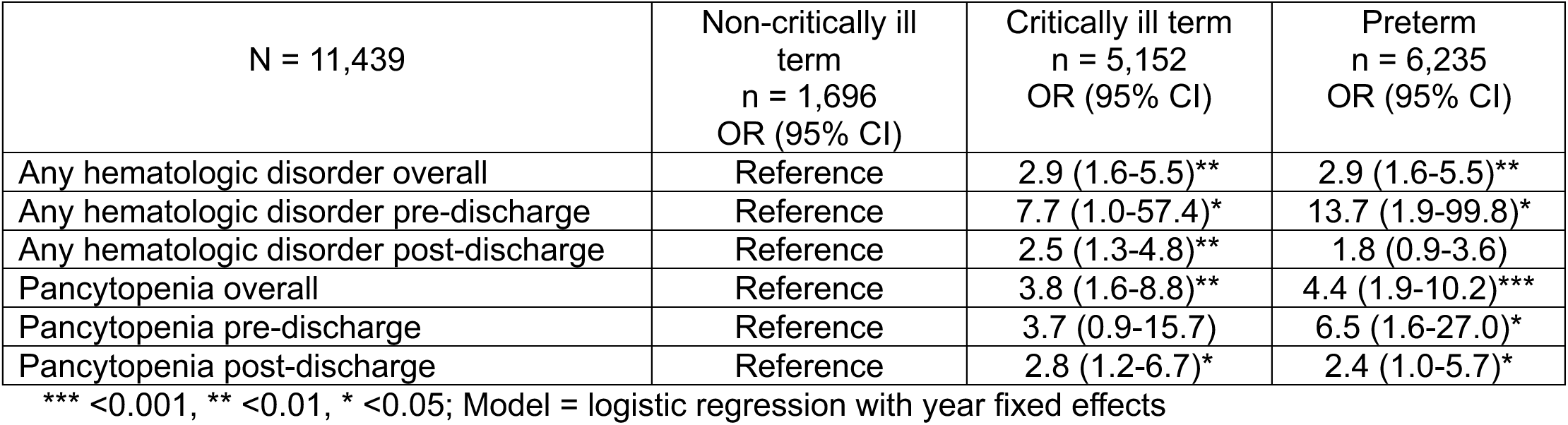
Logistic regression predicting any hematologic disorders or pancytopenia overall, pre-discharge, post-discharge.

| N = 11,439 | Non-critically ill term<br>n = 1,696<br>OR (95% CI) | Critically ill term<br>n = 5,152<br>OR (95% CI) | Preterm<br>n = 6,235<br>OR (95% CI) |
| --- | --- | --- | --- |
| Any hematologic disorder overall | Reference | 2.9 (1.6-5.5)** | 2.9 (1.6-5.5)** |
| Any hematologic disorder pre-discharge | Reference | 7.7 (1.0-57.4)* | 13.7 (1.9-99.8)* |
| Any hematologic disorder post-discharge | Reference | 2.5 (1.3-4.8)** | 1.8 (0.9-3.6) |
| Pancytopenia overall | Reference | 3.8 (1.6-8.8)** | 4.4 (1.9-10.2)*** |
| Pancytopenia pre-discharge | Reference | 3.7 (0.9-15.7) | 6.5 (1.6-27.0)* |
| Pancytopenia post-discharge | Reference | 2.8 (1.2-6.7)* | 2.4 (1.0-5.7)* |

\*\*\* <0.001, \*\* <0.01, \* <0.05; Model = logistic regression with year fixed effects

**Supplemental Table 5.**
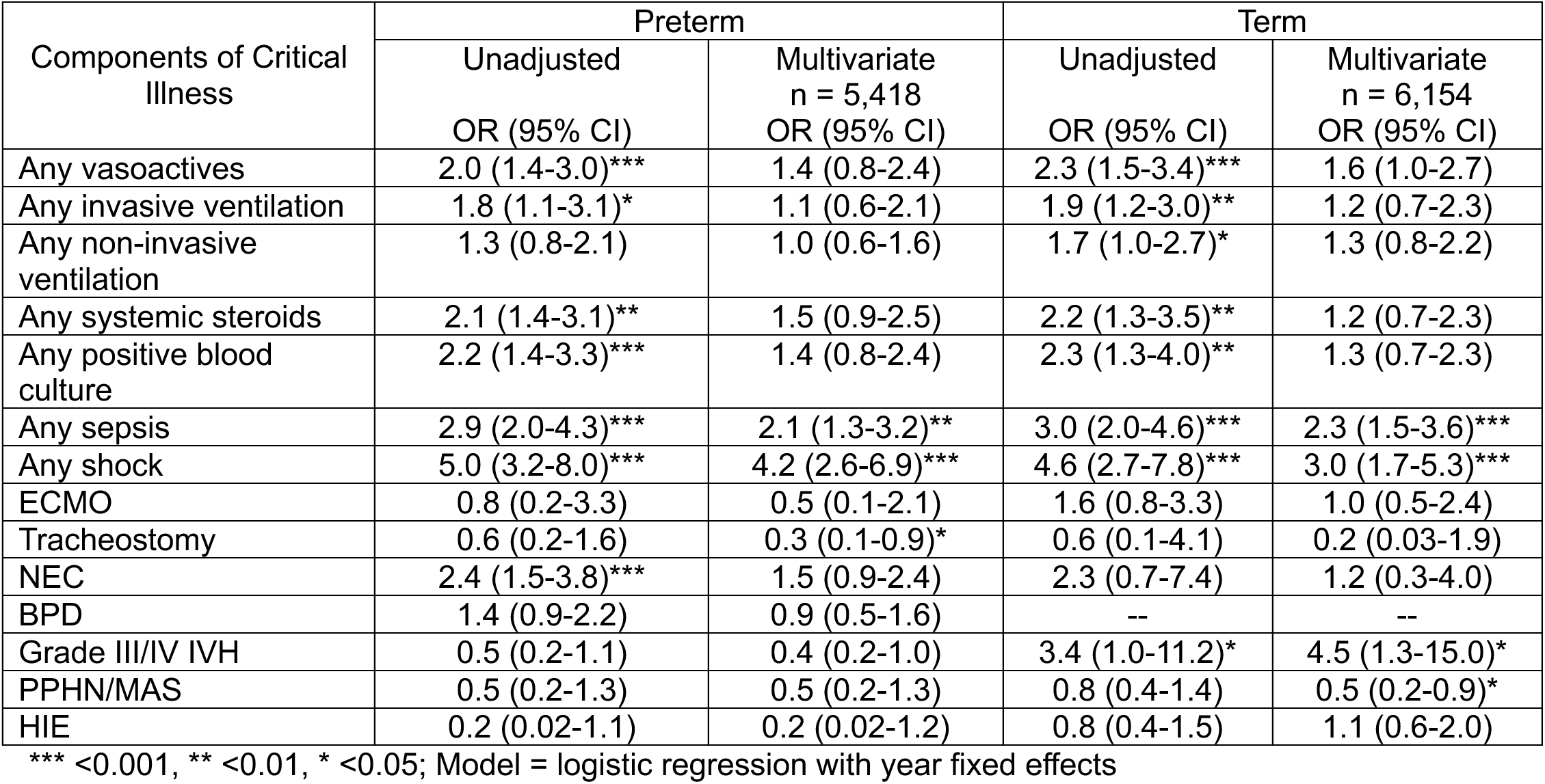
Logistic regression with components of critical illness predicting any hematologic disorder, stratified on prematurity.

| Components of Critical Illness | Preterm |  | Term |  |
| --- | --- | --- | --- | --- |
|  | Unadjusted<br>OR (95% CI) | Multivariate<br>n = 5,418<br>OR (95% CI) | Unadjusted<br>OR (95% CI) | Multivariate<br>n = 6,154<br>OR (95% CI) |
| Any vasoactives | 2.0 (1.4-3.0)*** | 1.4 (0.8-2.4) | 2.3 (1.5-3.4)*** | 1.6 (1.0-2.7) |
| Any invasive ventilation | 1.8 (1.1-3.1)* | 1.1 (0.6-2.1) | 1.9 (1.2-3.0)** | 1.2 (0.7-2.3) |
| Any non-invasive ventilation | 1.3 (0.8-2.1) | 1.0 (0.6-1.6) | 1.7 (1.0-2.7)* | 1.3 (0.8-2.2) |
| Any systemic steroids | 2.1 (1.4-3.1)** | 1.5 (0.9-2.5) | 2.2 (1.3-3.5)** | 1.2 (0.7-2.3) |
| Any positive blood culture | 2.2 (1.4-3.3)*** | 1.4 (0.8-2.4) | 2.3 (1.3-4.0)** | 1.3 (0.7-2.3) |
| Any sepsis | 2.9 (2.0-4.3)*** | 2.1 (1.3-3.2)** | 3.0 (2.0-4.6)*** | 2.3 (1.5-3.6)*** |
| Any shock | 5.0 (3.2-8.0)*** | 4.2 (2.6-6.9)*** | 4.6 (2.7-7.8)*** | 3.0 (1.7-5.3)*** |
| ECMO | 0.8 (0.2-3.3) | 0.5 (0.1-2.1) | 1.6 (0.8-3.3) | 1.0 (0.5-2.4) |
| Tracheostomy | 0.6 (0.2-1.6) | 0.3 (0.1-0.9)* | 0.6 (0.1-4.1) | 0.2 (0.03-1.9) |
| NEC | 2.4 (1.5-3.8)*** | 1.5 (0.9-2.4) | 2.3 (0.7-7.4) | 1.2 (0.3-4.0) |
| BPD | 1.4 (0.9-2.2) | 0.9 (0.5-1.6) | -- | -- |
| Grade III/IV IVH | 0.5 (0.2-1.1) | 0.4 (0.2-1.0) | 3.4 (1.0-11.2)* | 4.5 (1.3-15.0)* |
| PPHN/MAS | 0.5 (0.2-1.3) | 0.5 (0.2-1.3) | 0.8 (0.4-1.4) | 0.5 (0.2-0.9)* |
| HIE | 0.2 (0.02-1.1) | 0.2 (0.02-1.2) | 0.8 (0.4-1.5) | 1.1 (0.6-2.0) |

\*\*\* <0.001, \*\* <0.01, \* <0.05; Model = logistic regression with year fixed effects

**Supplemental Table 6.** Logistic regression with components of critical illness predicting pancytopenia, stratified on prematurity.

| Components of Critical Illness | Preterm |  | Term |  |
| --- | --- | --- | --- | --- |
|  | Unadjusted<br>OR (95% CI) | Multivariate<br>n = 5,418<br>OR (95% CI) | Unadjusted<br>OR (95% CI) | Multivariate<br>n = 6,162<br>OR (95% CI) |
| Any vasoactives | 2.3 (1.5-3.6)*** | 1.5 (0.8-2.6) | 2.7 (1.7-4.3)*** | 1.8 (1.0-3.2) |
| Any invasive ventilation | 1.8 (1.0-3.2)* | 1.0 (0.5-2.1) | 1.8 (1.0-3.1)* | 0.8 (0.4-1.7) |
| Any non-invasive ventilation | 1.4 (0.8-2.3) | 1.1 (0.6-1.8) | 2.1 (1.2-3.8)* | 1.6 (0.8-3.0) |
| Any systemic steroids | 2.2 (1.4-3.4)** | 1.4 (0.8-2.5) | 2.9 (1.7-5.0)*** | 1.7 (0.8-3.4) |
| Any positive blood culture | 2.7 (1.7-4.2)*** | 1.7 (1.0-3.0) | 3.1 (1.7-5.8)*** | 1.6 (0.8-3.2) |
| Any sepsis | 3.4 (2.2-5.1)*** | 2.1 (1.3-3.3)** | 3.6 (2.1-5.9)*** | 2.5 (1.5-4.3)** |
| Any shock | 6.2 (3.8-10.0)*** | 4.7 (2.8-8.0)*** | 4.9 (2.6-9.2)*** | 3.0 (1.6-5.9)** |
| ECMO | 1.1 (0.3-4.4) | 0.6 (0.1-2.5) | 2.3 (1.1-4.9)* | 1.4 (0.6-3.4) |
| Tracheostomy | 0.7 (0.3-2.0) | 0.4 (0.1-1.1) | -- | -- |
| NEC | 2.7 (1.6-4.4)*** | 1.6 (0.9-2.8) | 1.1 (0.1-7.9) | 0.5 (0.1-3.7) |
| BPD | 1.3 (0.8-2.1) | 0.8 (0.4-1.4) | -- | -- |
| Grade III/IV IVH | 0.4 (0.1-1.1) | 0.4 (0.1-1.0) | 1.5 (0.2-10.9) | 2.0 (0.3-15.1) |
| PPHN/MAS | 0.5 (0.2-1.5) | 0.5 (0.2-1.4) | 1.0 (0.5-1.8) | 0.5 (0.2-1.1) |
| HIE | 0.2 (0.03-1.4) | 0.2 (0.03-1.5) | 1.0 (0.5-1.9) | 1.4 (0.7-2.9) |

\*\*\* <0.001, \*\* <0.01, \* <0.05; Model = logistic regression with year fixed effects

**Supplemental Table 7.** Logistic regression with components of critical illness predicting post-discharge pancytopenia, stratified on prematurity.

| Components of Critical Illness | Preterm |  | Term |  |
| --- | --- | --- | --- | --- |
|  | Unadjusted<br>OR (95% CI) | Multivariate<br>n = 4,019<br>OR (95% CI) | Unadjusted<br>OR (95% CI) | Multivariate<br>n = 5,769<br>OR (95% CI) |
| Any vasoactives | 1.7 (0.9-3.2) | 1.4 (0.7-2.9) | 2.0 (1.2-3.6) * | 1.7 (0.9-3.3) |
| Any invasive ventilation | 1.6 (0.7-3.6) | 1.3 (0.5-3.2) | 1.7 (0.9-3.2) | 1.0 (0.4-2.3) |
| Any non-invasive ventilation | 1.6 (0.7-3.5) | 1.3 (0.6-3.0) | 1.5 (0.8-2.8) | 1.1 (0.5-2.3) |
| Any systemic steroids | 1.7 (0.9-3.3) | 1.2 (0.5-2.7) | 2.0 (1.0-3.9)* | 1.4 (0.6-3.3) |
| Any positive blood culture | 1.6 (0.8-3.2) | 1.0 (0.4-2.2) | 2.1 (1.0-4.7) | 1.4 (0.6-3.4) |
| Any sepsis | 3.8 (2.0-7.0)*** | 3.0 (1.5-6.0)** | 2.5 (1.4-4.5)** | 2.0 (1.0-3.7)* |
| Any shock | 10.8 (5.9-20.0)*** | 8.6 (4.4-16.6)*** | 5.0 (2.5-10.3)*** | 3.6 (1.7-7.7)** |
| ECMO | -- | -- | 1.8 (0.7-4.6) | 1.2 (0.4-3.6) |
| Tracheostomy | 1.0 (0.3-3.2) | 0.7 (0.2-2.6) | -- | -- |
| NEC | 1.0 (0.4-2.5) | 0.6 (0.2-1.7) | -- | -- |
| BPD | 0.7 (0.3-1.5) | 0.5 (0.2-1.4) | -- | -- |
| Grade III/IV IVH | 0.2 (0.03-1.5) | 0.2 (0.03-1.6) | 1.8 (0.2-13.7) | 2.2 (0.3-16.8) |
| PPHN/MAS | 0.3 (0.04-1.9) | 0.3 (0.03-1.9) | 0.9 (0.4-2.0) | 0.6 (0.2-1.4) |
| HIE | -- | -- | 1.5 (0.7-3.1) | 1.3 (0.6-2.9) |

\*\*\* <0.001, \*\* <0.01, \* <0.05; Model = logistic regression with year fixed effects

## References

1 Crump, C., Groves, A., Sundquist, J. & Sundquist, K. Association of Preterm Birth With Long-term Risk of Heart Failure Into Adulthood. JAMA Pediatr 175, 689–697 (2021). 10.1001/jamapediatrics.2021.0131

2 Kidokoro, H. et al. Brain injury and altered brain growth in preterm infants: predictors and prognosis. Pediatrics 134, e444–453 (2014). 10.1542/peds.2013-2336

3 Markopoulou, P., Papanikolaou, E., Analytis, A., Zoumakis, E. & Siahanidou, T. Preterm Birth as a Risk Factor for Metabolic Syndrome and Cardiovascular Disease in Adult Life: A Systematic Review and Meta-Analysis. J Pediatr 210, 69–80 e65 (2019). 10.1016/j.jpeds.2019.02.041

4 Pulakka, A. et al. Preterm birth and asthma and COPD in adulthood: a nationwide register study from two Nordic countries. Eur Respir J 61 (2023). 10.1183/13993003.01763-2022

5 Sanchez-Solano, N. J. et al. Lung Function in a Multiethnic U.S. Cohort of Adolescents and Adults Born Preterm in the New BPD Era. Pediatr Pulmonol 60, e71226 (2025). 10.1002/ppul.71226

6 Saigal, S. et al. Growth trajectories of extremely low birth weight infants from birth to young adulthood: a longitudinal, population-based study. Pediatr Res 60, 751–758 (2006). 10.1203/01.pdr.0000246201.93662.8e

7 Horvath-Puho, E. et al. Mortality, neurodevelopmental impairments, and economic outcomes after invasive group B streptococcal disease in early infancy in Denmark and the Netherlands: a national matched cohort study. Lancet Child Adolesc Health 5, 398–407 (2021). 10.1016/S2352-4642(21)00022-5

8 Ijsselstijn, H. & van Heijst, A. F. Long-term outcome of children treated with neonatal extracorporeal membrane oxygenation: increasing problems with increasing age. Semin Perinatol 38, 114–121 (2014). 10.1053/j.semperi.2013.11.009

9 Thom, C. S. et al. Extreme thrombocytosis is associated with critical illness and young age, but not increased thrombotic risk, in hospitalized pediatric patients. J Thromb Haemost 18, 3352–3358 (2020). 10.1111/jth.15103

10 Widness, J. A. Pathophysiology of Anemia During the Neonatal Period, Including Anemia of Prematurity. Neoreviews 9, e520 (2008). 10.1542/neo.9-11-e520

11 Christensen, R. D. et al. Thrombocytopenia among extremely low birth weight neonates: data from a multihospital healthcare system. J Perinatol 26, 348–353 (2006). 10.1038/sj.jp.7211509

12 Maheshwari, A. Neutropenia in the newborn. Curr Opin Hematol 21, 43–49 (2014). 10.1097/MOH.0000000000000010

13 Deschmann, E. et al. Clinical Practice Guideline for Red Blood Cell Transfusion Thresholds in Very Preterm Neonates. JAMA Netw Open 7, e2417431 (2024). 10.1001/jamanetworkopen.2024.17431

14 Baumann, A. M. & Ellegast, J. M. Inflammatory signaling in the pathogenesis of acute myeloid leukemia. Hemasphere 9, e70188 (2025). 10.1002/hem3.70188

15 Kapadia, C. D. et al. Clonal dynamics and somatic evolution of haematopoiesis in mouse. Nature 641, 681–689 (2025). 10.1038/s41586-025-08625-8

16 Huang, Q. T., Gao, Y. F., Zhong, M. & Yu, Y. H. Preterm Birth and Subsequent Risk of Acute Childhood Leukemia: a Meta-Analysis of Observational Studies. Cell Physiol Biochem 39, 1229–1238 (2016). 10.1159/000447828

17 Seppala, L. K., Vettenranta, K., Leinonen, M. K., Tommiska, V. & Madanat-Harjuoja, L. M. Preterm birth, neonatal therapies and the risk of childhood cancer. Int J Cancer 148, 2139–2147 (2021). 10.1002/ijc.33376

18 Schuermans, A. et al. Birth Weight Is Associated With Clonal Hematopoiesis of Indeterminate Potential and Cardiovascular Outcomes in Adulthood. J Am Heart Assoc 12, e030220 (2023). 10.1161/JAHA.123.030220

19 Chen, C. C. et al. Association between neonatal sepsis and lung function in school-age children born preterm. Pediatr Res (2026). 10.1038/s41390-026-04931-7

20 Glaser, K. et al. Neonatal Sepsis Episodes and Retinopathy of Prematurity in Very Preterm Infants. JAMA Netw Open 7, e2423933 (2024). 10.1001/jamanetworkopen.2024.23933

21 Mansell, T. et al. Early life infection and proinflammatory, atherogenic metabolomic and lipidomic profiles in infancy: a population-based cohort study. Elife 11 (2022). 10.7554/eLife.75170

22 Olin, A. et al. Stereotypic Immune System Development in Newborn Children. Cell 174, 1277–1292 e1214 (2018). 10.1016/j.cell.2018.06.045

23 Biswas, N. et al. Survivors of polymicrobial sepsis are refractory to G-CSF-induced emergency myelopoiesis and hematopoietic stem and progenitor cell mobilization. Stem Cell Reports 19, 639–653 (2024). 10.1016/j.stemcr.2024.03.007

24 de Laval, B. et al. C/EBPbeta-Dependent Epigenetic Memory Induces Trained Immunity in Hematopoietic Stem Cells. Cell Stem Cell 30, 112 (2023). 10.1016/j.stem.2022.12.005

25 Tran, B. T., Jeyanathan, V., Cao, R., Kaufmann, E. & King, K. Y. Hematopoietic stem and progenitor cells as a reservoir for trained immunity. Elife 14 (2025). 10.7554/eLife.106610

26 Zeng, A. G. X. et al. Human haematopoietic stem cells remember inflammatory stress. Nature (2026). 10.1038/s41586-026-10522-7

27 Swann, J. W. et al. Inflammation perturbs hematopoiesis by remodeling specific compartments of the bone marrow niche. Blood 147, 739–754 (2026). 10.1182/blood.2025029513

28 Cain, T. L., Derecka, M. & McKinney-Freeman, S. The role of the haematopoietic stem cell niche in development and ageing. Nat Rev Mol Cell Biol 26, 32–50 (2025). 10.1038/s41580-024-00770-8

29 Dulmovits, B. M. et al. A single cell atlas defines perinatal factors that drive murine bone marrow development. bioRxiv (2026). 10.64898/2026.01.21.700817

30 Liu, Y. et al. Dopamine signaling regulates hematopoietic stem and progenitor cell function. Blood 138, 2051–2065 (2021). 10.1182/blood.2020010419

31 Leisman, D. E. et al. Angiotensin II enhances bacterial clearance via myeloid signaling in a murine sepsis model. Proc Natl Acad Sci U S A 119, e2211370119 (2022). 10.1073/pnas.2211370119

32 Agha, N. H. et al. Vigorous exercise mobilizes CD34+ hematopoietic stem cells to peripheral blood via the beta(2)-adrenergic receptor. Brain Behav Immun 68, 66–75 (2018). 10.1016/j.bbi.2017.10.001

33 Maryanovich, M., Takeishi, S. & Frenette, P. S. Neural Regulation of Bone and Bone Marrow. Cold Spring Harb Perspect Med 8 (2018). 10.1101/cshperspect.a031344

34 Hartung, H. D., Olson, T. S. & Bessler, M. Acquired aplastic anemia in children. Pediatr Clin North Am 60, 1311–1336 (2013). 10.1016/j.pcl.2013.08.011

35 Siegel, R. L., Kratzer, T. B., Giaquinto, A. N., Sung, H. & Jemal, A. Cancer statistics, 2025. CA Cancer J Clin 75, 10–45 (2025). 10.3322/caac.21871

